# Wastewater as a back door to serology?

**DOI:** 10.1101/2022.11.11.22282224

**Authors:** Marie LittleFawn Agan, William R. Taylor, Isaiah Young, William A. Willis, Gari D. New, Halee Lair, Anastasia Murphy, Anna Marinelli, Md Ariful Islam Juel, Mariya Munir, Alex Dornburg, Jessica Schlueter, Cynthia Gibas

## Abstract

Wastewater surveillance is a powerful tool for monitoring the prevalence of infectious disease. Systems for wastewater monitoring were put in place throughout the world during the COVID-19 pandemic. These systems use viral RNA copies as the basis of estimates of COVID-19 cases in the sewershed area, thereby providing data critical for public health responses. However, the potential to measure other biomarkers in wastewater during outbreaks has not been fully explored. Here we report a novel approach for detecting specific human antibodies from wastewater. We measured the abundance of anti-SARS-CoV-2 spike IgG and IgA from fresh samples of community wastewater and from archived frozen samples dating from 2020-22. The assay described can be performed with readily available reagents, at a moderate per-sample cost. Our findings demonstrate the feasibility of noninvasive serological surveillance via wastewater, enabling a new approach to immunity-based monitoring of populations.

---

Seroprevalence surveillance plays a critical role in detection and epidemiology of infectious disease outbreaks.^1,2^ Serological evidence of infection can also be used as an indicator of likelihood of zoonotic spillover and precursor to emergence of novel diseases.^3^ The onset of the COVID-19 pandemic led to a call for a Global Immunological Observatory^4^ that would provide information about susceptibility of populations based on aggregate serological data, allowing for public health action in advance of waves of infection. However, there are several requirements which are barriers to large-scale data collection and the achievement of such an aim. One of these is the need to collect large numbers of serological samples from individual patients, which implies the need for patient participation, or to reliably obtain material from clinical discards. Such requirements are especially challenging in low and middle income countries^5^ but can pose a challenge even in resource-rich areas. In the case of the SARS-CoV-2 virus, a massive amount of genomic surveillance data was collected as governments required testing and made free testing available, but as pandemic mitigation measures have declined or ended, clinical test discards for sequencing have become difficult to obtain and surveillance by genomic sequencing has declined significantly. In turn, health departments have turned to sequencing and viral variant identification out of wastewater as a non-invasive method that can be conducted independent of patient choices about treatment and testing. The widespread adaptation of wastewater surveillance raises a question: can wastewater be used as a source for other biomarkers that can illuminate global immunological trends?

Wastewater surveillance has proven useful at both the regional scale for detection of SARS-CoV-2 infection trends^6–8^, and at the local scale for guiding rapid response to emerging outbreaks.^9,10^ The main modes of wastewater surveillance during the COVID-19 pandemic have been quantitative detection and variant sequencing of viral RNA isolated from wastewater. However, the potential for monitoring other immunologically relevant molecules in wastewater has not been as extensively explored. Human antibodies against SARS-CoV-2 offer a particularly promising target. Neutralization activity against SARS-CoV-2 is directly correlated with levels of anti-SARS-CoV-2 IgG or IgA antibodies^11^. Short-term longitudinal studies of SARS-CoV-2 neutralizing antibodies in naturally infected, vaccinated, or boosted individuals have made clear that antibody protection against future infection wanes within a period of months^12,13^. As governmental interventions ebb globally and rates of booster vaccine uptake continue to decline, there is little doubt that COVID-19 will remain common in future years, as an endemic disease^14^. Understanding the susceptibility of populations to future waves of infections is critical for mitigating an endemic state that results in unacceptable morbidity and mortality. Expanding the scope of surveillance to include detection of antibody levels would offer a proactive approach that can identify populations at risk of future infection.

Immunoassay systems have been widely used as a biotechnology tool for detection of SARS-CoV-2 RNA^15,16^ and viral antigens in clinical samples. Samples for immunoassays are typically collected from serum^17^, but there have also been recent advances in use of immunosensors targeting the viral antigen protein in wastewater.^18,19^ Human antibodies shed in urine and feces have not been targeted as an immune biomarker in wastewater, despite three years of intense focus on monitoring and mitigation of COVID-19 infections. Antibody measurements from fecal samples have already been highlighted as a non-invasive means to rapidly monitor the immune status of wildlife^20^ Similarly, measurements from fecal samples are increasingly being used for studying various aspects of human health,^21,22^ as fecal antibody levels are correlated with antibody levels in serum^23^, and have been demonstrated as a potential biomarker for detection of viral^24^ and parasitic infections^25^ in human urine samples. This raises the question of whether similar human antibody data can be extracted from wastewater.

Samples from the COVID-19 pandemic offer an ideal case study for developing approaches to extract antibody data from wastewater. Infection by SARS-CoV-2 elicits the secretion of mucosal IgG and IgA antibodies.^26^ The IgA response waxes and wanes rapidly, being detectable in serum within days of infection, before falling below the limit of detection within weeks. In contrast, the signal of the IgG response in serum rises more slowly and may persist for months^27^, potentially remaining above the limit of detection even longer (**Figure 1**). Coronavirus-specific antibodies are also known to be recoverable from fecal samples in animal models^28^, which suggests the possibility of human anti-SARS-CoV-2 antibody recovery from wastewater. Antibodies developed against SARS-CoV-2 are among the most well studied in immunology, with numerous approaches already developed for detecting specific antibodies that can be leveraged for application in wastewater.

**Figure 1.**
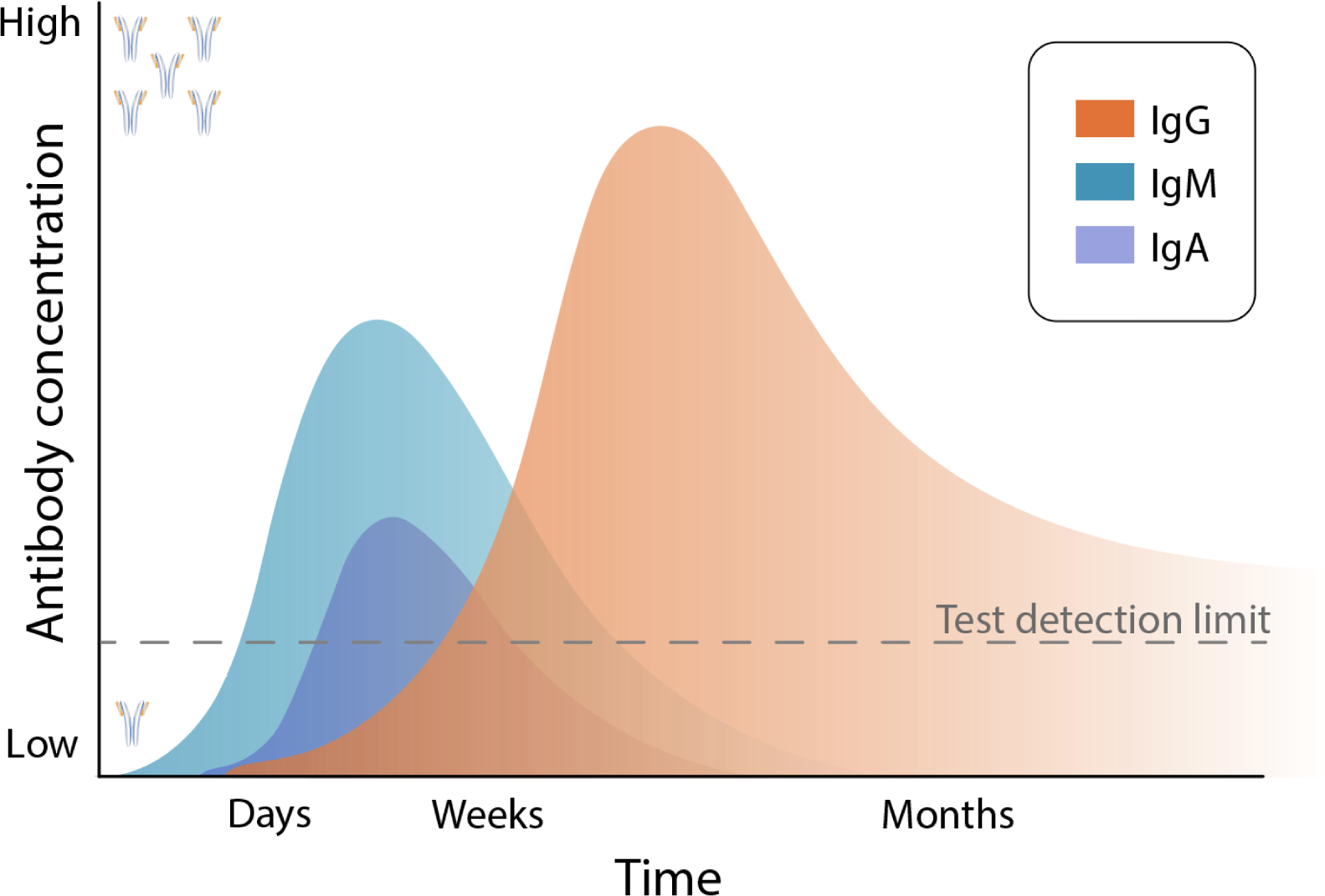
Generalized timeline of antibody waxing and waning in the human immune response.

Here we present a protocol for an enzyme-linked immunosorbent assay (ELISA) to detect anti-spike (S) and anti-nucleocapsid (N) SARS-CoV-2 IgG and IgA antibodies from wastewater. We assess how detection varies by dilution and demonstrate the functionality of our approach on both freshly collected and archived frozen samples. This protocol enables new research directions, future monitoring, retroactive studies of archived samples collected during the SARS-CoV-2 pandemic through the ability to recover antibody signals from archived frozen wastewater. The ability to retroactively recover antibody signals from archived samples is of particular importance, as SARS-CoV-2 RNA that is currently used as an indicator is not durable in frozen wastewater samples, even those held at −80°C.^29^ By monitoring specific antibody levels in a sewershed, our approach can be used to reveal waning of a durable immune response in the population after episodes of widespread infection or after seasonal vaccination campaigns. This approach complements existing wastewater-based viral detection and sequencing efforts, increasing the scope of wastewater surveillance to provide data critical for public health policy and decision-making.

## Results

### Detection of bulk human IgG and IgA in fresh wastewater samples

We initially assayed whether any antibodies were detectable in fresh wastewater by measuring the abundance of total human IgG (Figure 2a) and IgA (Figure 2b) in samples dating from October 2022. After concentration, bulk human IgG and IgA were both detected at concentrations that exceeded the upper detection limit of the ELISA assay, at all dilutions tested. Exactly as in classic serology, our approach yields ELISA assays displaying a color change in the positive sample that is visible to the eye (Figure 2a,b), and can also be quantified as the optical density at 450nm (OD450) using a plate reader (Table 1a,b). In the case of this example, the lower detection limit used in this experiment was 2x the negative control signal, as recommended by the manufacturer. For IgG, all dilutions of the neat sample by less than 1:16 gave a signal above background (Table 1a) and for IgA, all dilutions by less than 1:32 gave a signal above background by the manufacturer’s definition. These OD450 readings correspond to 5.08 ng/ml of IgG and 1.52 ng/ml of IgA in concentrated wastewater. For comparison, serum levels of total human IgG and IgA are known to be 8-18 mg/ml and 0.4 to 2.2 mg/ml respectively, or approximately 1000-fold greater.^30^

**Figure 2a.**
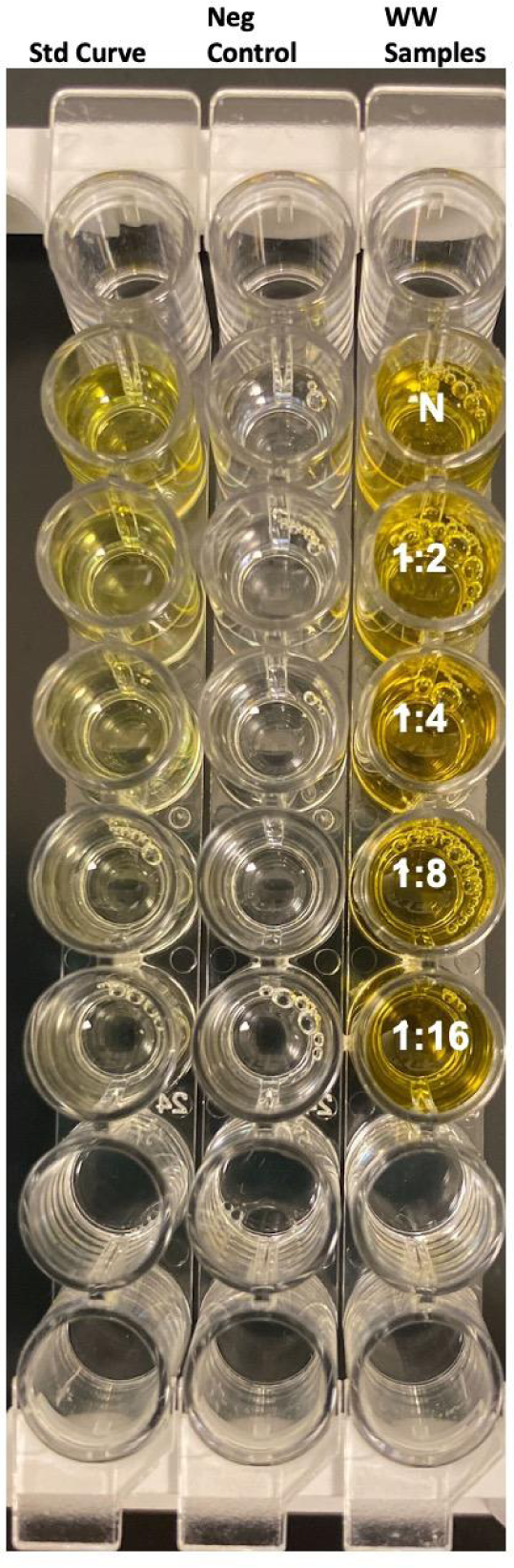
Representative ELISA plate assay, for concentrated wastewater evaluated for total IgG by ELISA. The first column of wells in the plate contains a standard reference dilution series. The second column contains the negative control. Wastewater samples are labeled by row as Neat or by dilution factor.

**Figure 2b.**
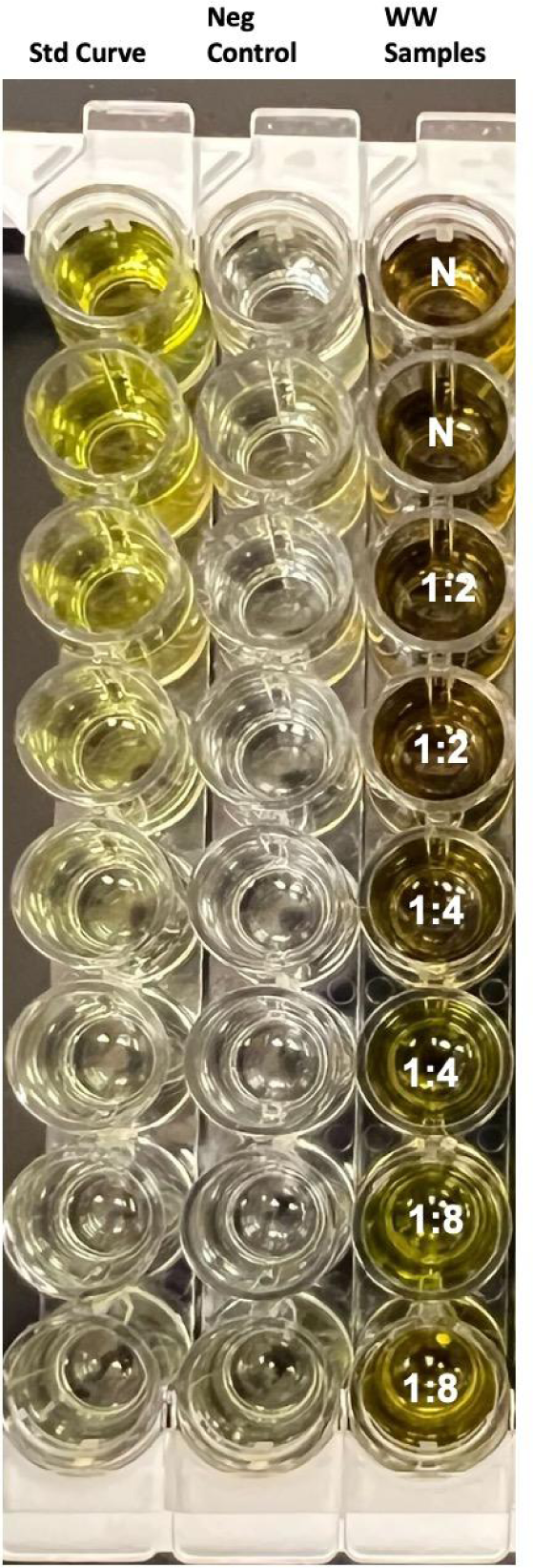
Representative ELISA plate assay for concentrated wastewater evaluated for total IgA by ELISA. The first column of wells in the plate contains a standard reference dilution series. The second column contains the negative control. Wastewater samples are labeled by row as Neat or by dilution factor.

**Table 1a.**
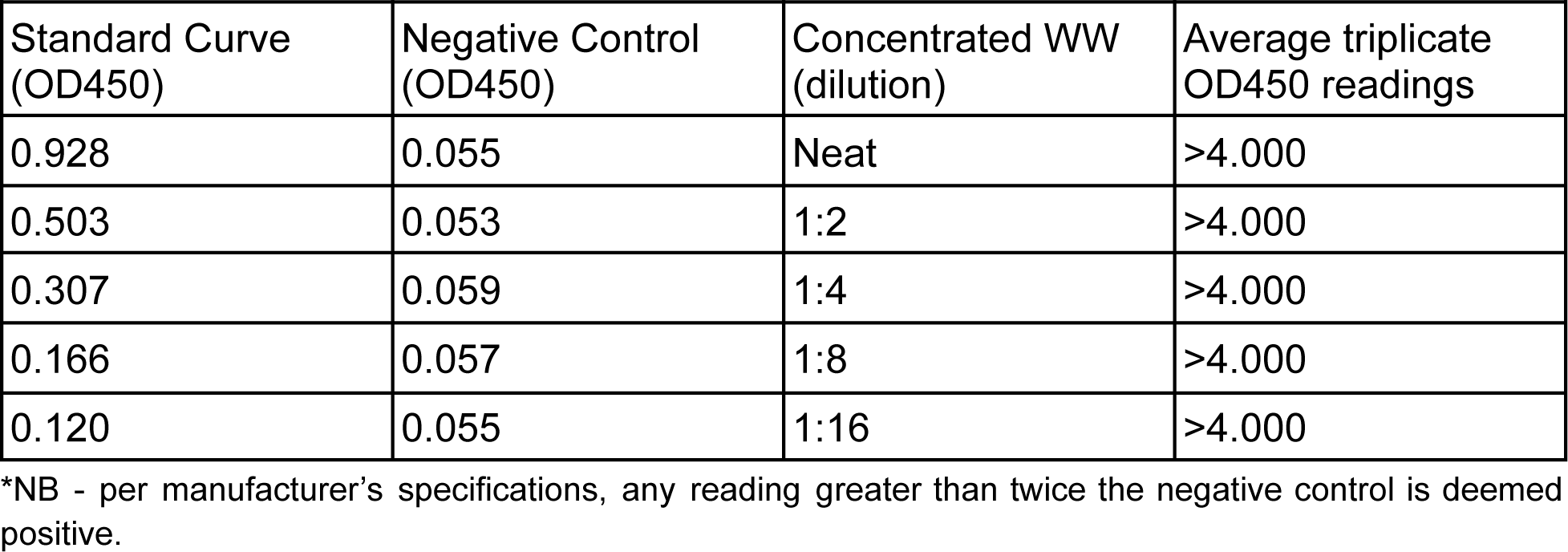
OD450 readings of total human IgG on wastewater.

| Standard Curve (OD450) | Negative Control (OD450) | Concentrated WW (dilution) | Average triplicate OD450 readings |
| --- | --- | --- | --- |
| 0.928 | 0.055 | Neat | >4.000 |
| 0.503 | 0.053 | 1:2 | >4.000 |
| 0.307 | 0.059 | 1:4 | >4.000 |
| 0.166 | 0.057 | 1:8 | >4.000 |
| 0.120 | 0.055 | 1:16 | >4.000 |

\*NB - per manufacturer's specifications, any reading greater than twice the negative control is deemed positive.

**Table 1b.** OD450 readings of total human IgA on wastewater.*

| Standard Curve (OD450) | Negative Control (OD450) | Concentrated WW (dilution) | Average triplicate OD450 readings |
| --- | --- | --- | --- |
| 2.611 | 0.124 | Neat | >4.000 |
| 1.577 | 0.227 | 1:2 | >4.000 |
| 0.873 | 0.108 | 1:4 | >4.000 |
| 0.485 | 0.099 | 1:8 | >4.000 |
| 0.297 | 0.1 | 1:16 | >4.000 |
| 0.197 | 0.103 | 1:32 | >4.000 |

\*NB - per manufacturer's specifications, any reading greater than twice the negative control is deemed positive.

### Detection of anti-SARS-CoV-2 spike protein antibodies

Given the ability to detect the presence of bulk human IgG and IgA, we next assayed specific human anti-SARS-CoV-2 spike (S) protein IgG and IgA in concentrated samples. We used both fresh samples, and frozen samples from two timepoints in an archived longitudinal series collected during the SARS-CoV-2 pandemic. We found that both anti-S IgG (Figure 3, Table 2) and anti-S IgA antibodies were detectable in quantities within or over the range of detection of the ELISA assay. This was true for fresh samples and for frozen samples. In general, anti-S IgA was found to be present in higher abundance than anti-S IgG in recent fresh wastewater samples (Figure 4, Table 3). However, this was not always the case. To assess whether anti-S antibodies were detectable during the onset of the pandemic in North Carolina, we additionally assayed samples collected in June of 2020. During this time, Mecklenburg County NC had only experienced a cumulative 9294 known COVID-19 cases in a population of 1.12 million. In this case, we do not have a sufficient signal of anti-S IgG or anti-S IgA to be classified as positive. (Figures 3,4) Archival samples from the June 2020 time period do, however, still produce a very high generic human IgG signal (Figures S2-3, Tables S2-3), demonstrating that the lack of anti-S signal in June 2020 is due to its absence and not due to degradation of protein in the sample.

**Fig. 3.**
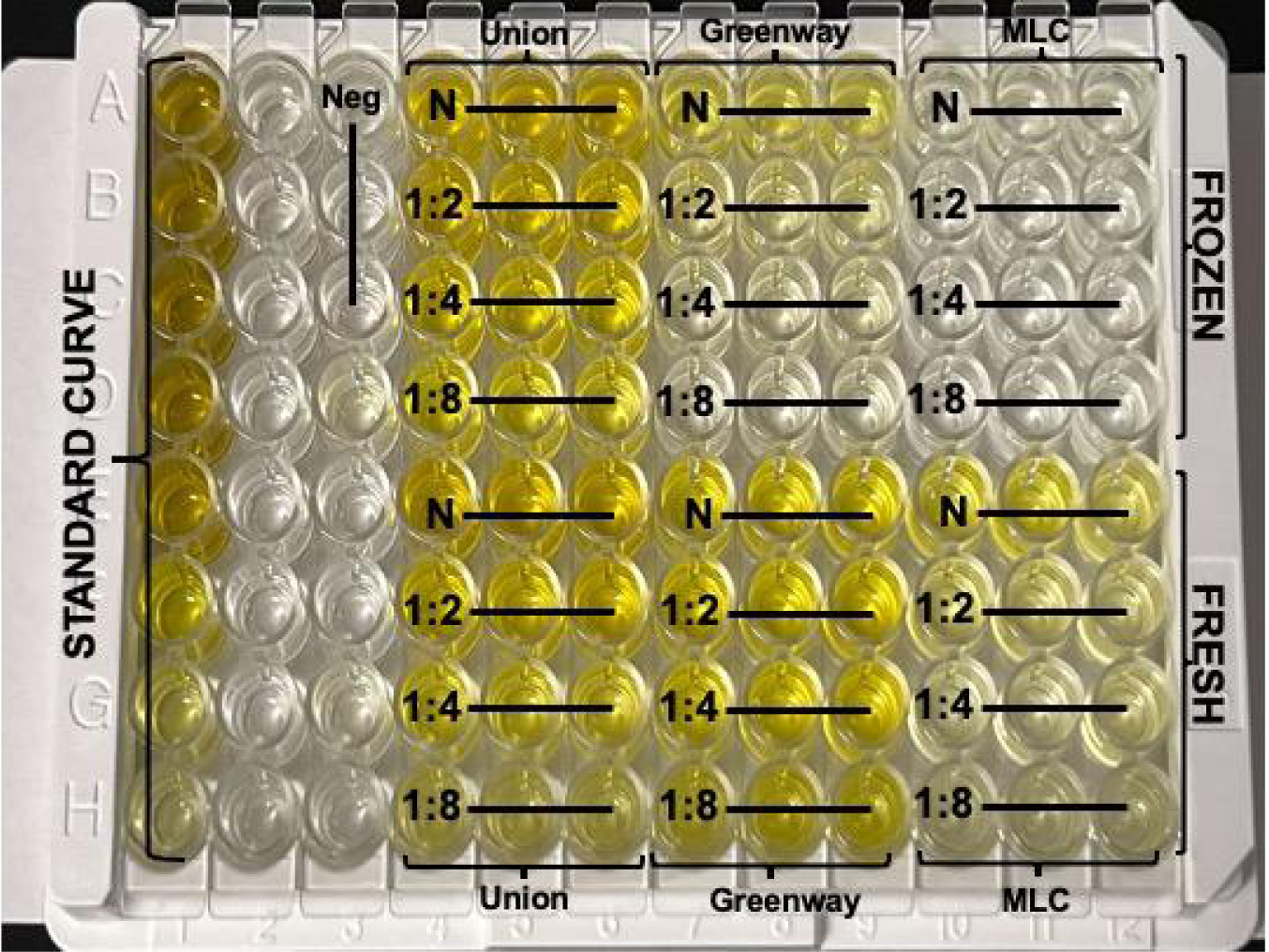
Concentrated wastewater evaluated for anti-SARS-Cov-2 S-Protein IgG by ELISA.

**Fig. 4.**
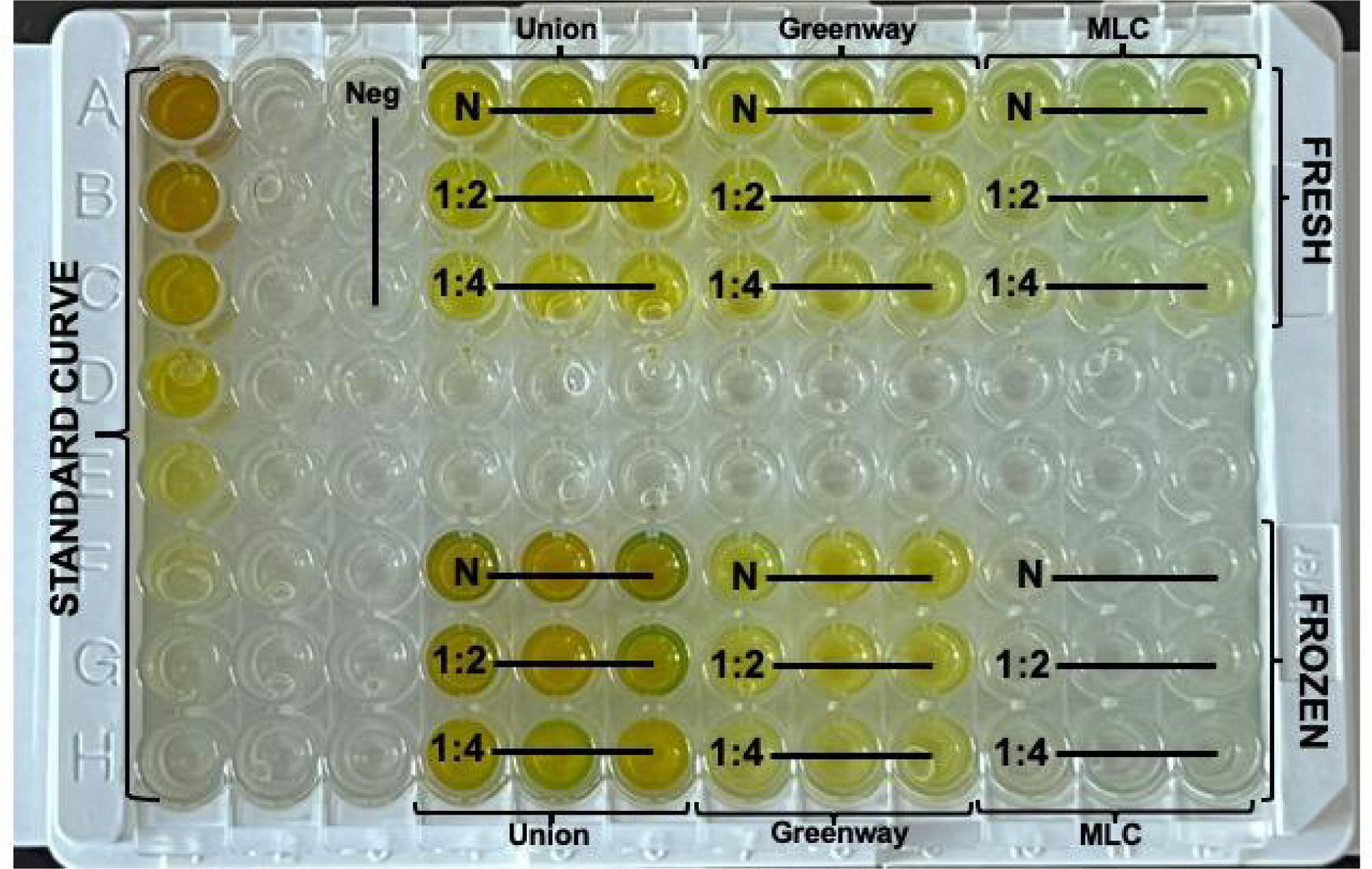
Concentrated wastewater evaluated for anti-SARS-Cov-2 S-Protein IgA by ELISA.

**Table 2.** Average triplicate OD450 readings for COVID-19 S-Protein (S1RBD) Human IgG.*

|  | Union<br>Fresh<br>10/2022 | Union<br>Frozen<br>2/2022 | Greenway<br>Fresh<br>10/2022 | Greenway<br>Frozen<br>2/2022 | MLC Fresh<br>10/2022 | MLC<br>Frozen<br>6/2020 |
| --- | --- | --- | --- | --- | --- | --- |
| neat | 2.58 | 2.36 | 0.971 | 0.519 | 0.599 | 0.088 |
| 1:2 | 1.42 | 2.04 | 1.09 | 0.223 | 0.291 | 0.060 |
| 1:4 | 0.705 | 1.21 | 0.972 | 0.135 | 0.189 | 0.061 |
| 1:8 | 0.391 | 0.722 | 0.695 | 0.098 | 0.163 | 0.059 |

\*NB - per manufacturer's specifications, any value greater than twice the negative control is deemed positive. Average Negative Control OD450: 0.053

**Table 3:** Average triplicate OD450 readings for COVID-19 S-Protein (S1RBD) Human IgA.

|  | Union<br>Fresh<br>10/2022 | Union<br>Frozen<br>2/2022 | Greenway<br>Fresh<br>10/2022 | Greenway<br>Frozen<br>2/2022 | MLC Fresh<br>10/2022 | MLC<br>Frozen<br>6/2020 |
| --- | --- | --- | --- | --- | --- | --- |
| neat | 2.236 | >4.000 | 0.952 | 0.865 | 0.475 | 0.107 |
| 1:2 | 1.632 | 3.772 | 0.751 | 0.704 | 0.401 | 0.103 |
| 1:4 | 0.973 | 2.889 | 0.601 | 0.516 | 0.268 | 0.1 |

\*NB - per manufacturer's specifications, any value greater than twice the negative control is deemed positive. Average Negative Control OD450: 0.091

### Assessing longitudinal trends of antibody concentration in wastewater

To assess longitudinal trends in antibody concentration, we applied our approach to wastewater collected at the UNC Charlotte Student Union at monthly intervals between April and December 2021. By April 2021, vaccination campaigns had begun and traffic on campus was increasing, and by August of 2021, the on-campus resident population had been returned to normal density. We measured anti-S IgG for each sample and also extended our approach to capture anti-SARS-CoV-2 Nucleocapsid (N) IgG. Comparing the abundance of anti-S to anti-N IgG reveals parallel temporal peaks of both antibodies in May of 2021 (Figure 6). However, there is a mismatch between the abundances anti-S IgG and anti-N IgG in September. When contrasted to the number of known on-campus positive COVID-19 tests for each month, anti-N IgG mirrors patterns of incidence (Figure 6a) while anti-S IgG does not (Figure 6b).

**Figure 5:**
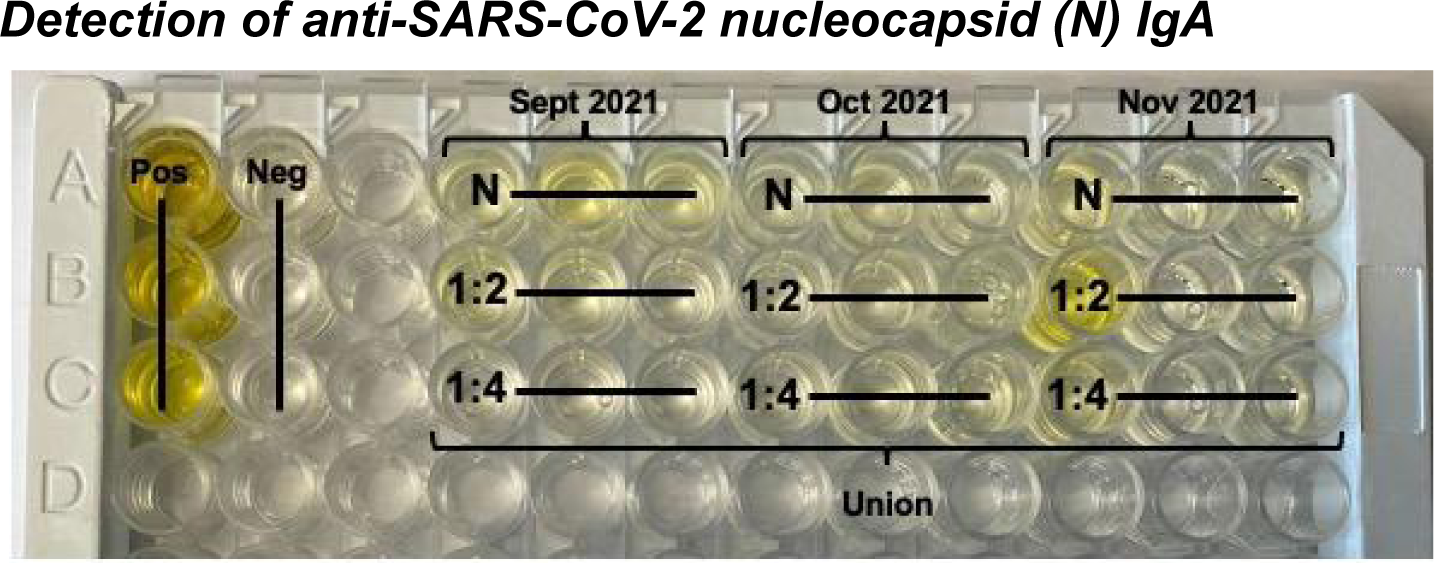
Concentrated wastewater evaluated for anti-SARS-CoV-2 N protein IgA by Qualitative ELISA.

**Figure 6:**
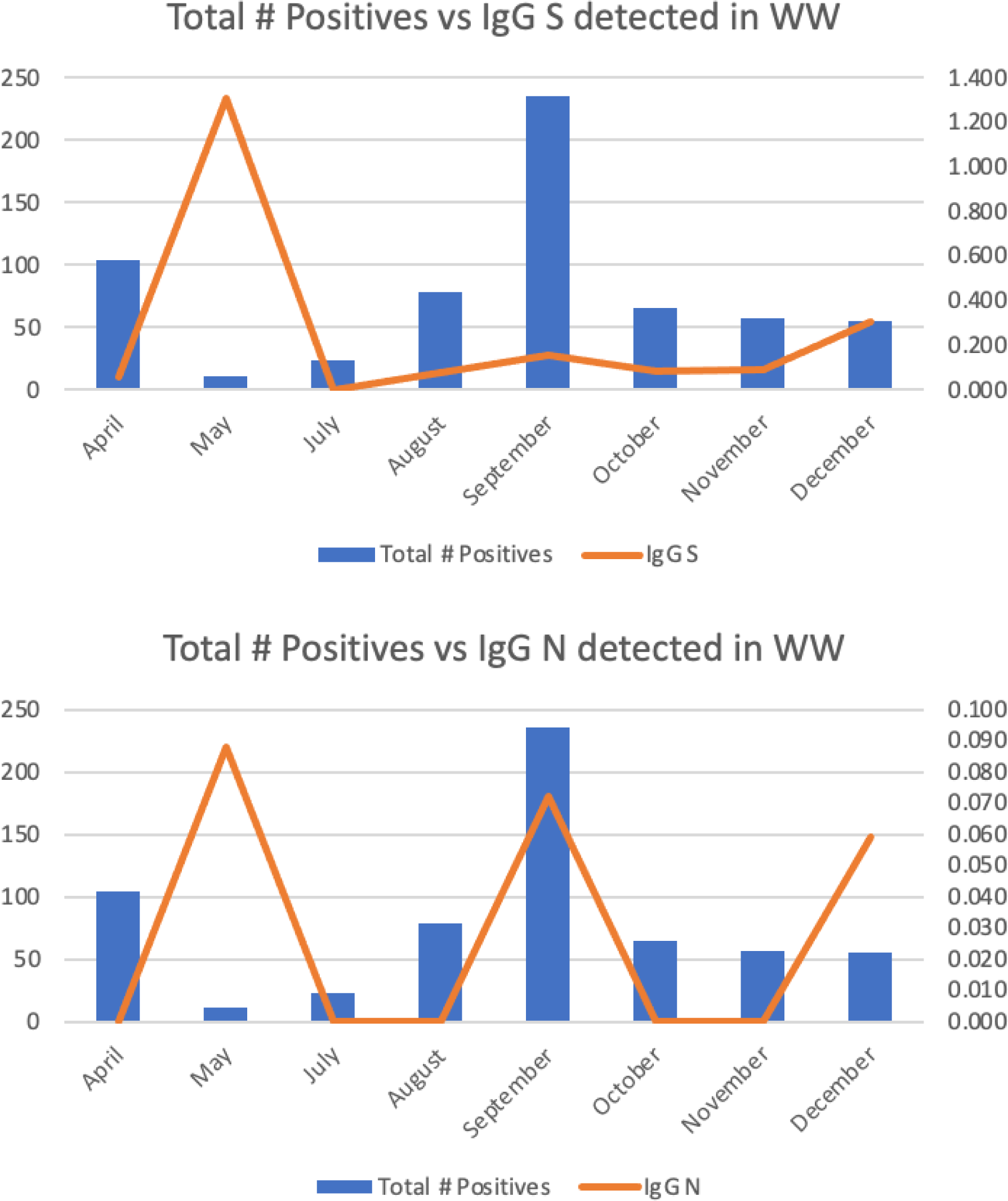
Anti-S and anti-N IgG timing in wastewater collected at the UNC-Charlotte Student Union from April to December 2021.

## Discussion

We have developed a means to quantify anti-SARS-CoV-2 antibody measurements from wastewater. We demonstrate the ability of our approach to detect specific human IgA and IgG antibodies against the SARS-CoV-2 spike (S) and nucleocapsid (N) proteins. Using our approach we quantified antibody levels from both fresh and archived frozen wastewater samples, facilitating future and retroactive studies of shed antibodies. We also observed a seasonal variation in antibody concentration that appears correlated with pandemic waves in an initial low-resolution time series. Our approach enables the development of a suite of novel applications for wastewater surveillance, such as monitoring changes in the immune status of populations, and the creation of quantitative models linking wastewater antibody concentrations to probabilistic models of seroprevalence.

It may be expected that detection and measurement of antibody levels would require concentration of very large volumes of wastewater, application of specialized processing methods, or that antibodies might rapidly degrade in wastewater environments to undetectable levels. However, this is not the case. We have demonstrated that human IgG and IgA are present in community wastewater in abundance and are easily detectable in samples concentrated from 30-60mL of raw wastewater, even in samples held at −80C for more than three years. Since the appearance of this work in preprint form in December 2022, the high abundance of human antibody signal in wastewater has been verified independently by Carrascal et al.^31^ using GC-MS based proteomic methods We demonstrate that specific SARS-CoV-2 antibodies can be detected with readily available equipment using a relatively inexpensive protocol. The cost of the reagents used amounts to $193/sample with ELISA kits supplied by Abcam and $166/sample with Raybio, using 3 replicates per sample. Controls are $130/run with Abcam and $40/run with Raybio. Although the current cost of ddPCR tests for viruses in wastewater is approximately $90/sample, an ELISA assay requires only a commonly available lab centrifuge and a plate reader, rather than more specialized instruments such as those used for ddPCR or GC-MS. The commercial ELISA assays used are validated to be resistant to detection of cross-reactive antibodies from other mammals, as well as to antibodies specific to other common human-infecting coronaviruses. Manufacturer-reported sensitivity of the assay is 94.7%, and specificity is 98.6%.^32^ Similar commercial ELISA assays are available for specific COVID-19 variants of concern such as delta and omicron, as well as other seasonally circulating viruses such as RSV and influenza, which have posed a renewed public health concern ^33^ after their temporary suppression ^34^ during the acute phase of the COVID-19 pandemic. With such a cost-efficient and reliable assay, it now becomes feasible to perform the investigations needed to validate detected antibodies in wastewater as a public health indicator.

Current wastewater surveillance approaches for viruses such as SARS-CoV-2 focus on measurements of viral abundance that reflect viral shedding by already-infected individuals. Such data is invaluable for identifying infection outbreaks and informing the needs of immediate public health responses.^35–37^ As antibody levels remain high weeks to months after infection ^38^, the continual detection of antibodies alone is not likely to be a good leading indicator for an impending local outbreak. However, antibody levels detected in wastewater could still have substantial utility for proactive public health planning and actions. For example, antibody detection could enable agencies to detect and address a community-wide waning of SARS-CoV-2 immunity with vaccination campaigns, before infections become common. Because our assay can distinguish anti-nucleocapsid (N) and anti-S antibodies, it would enable differentiation of recent circulating infection from waning of vaccine-mediated immunity. This is possible with the SARS-CoV-2 virus because commonly-used vaccines such as mRNA-1273 and BNT1262b currently elicit an immune response to the SARS-CoV-2 spike protein in isolation.^39^ In contrast, natural infection with SARS-CoV-2 also elicits immune response to other components of the virus.^39^ Therefore, the absence of anti-N paired with high anti-S levels would reveal a signature of a durable vaccine response, while the absence of both would reveal a population that is potentially at risk for future infection.

Wastewater samples by nature aggregate antibodies from a few to hundreds or thousands of individuals, which poses a challenge for direct interpretation of antibody levels detected and immune status. This opens the door to the development of new approaches that generalize the immune status of a population. For example, COVID-19 wastewater surveillance projects that submit to CDC’s National Wastewater Surveillance System^40^ gather site metadata describing water flow rates and sewershed populations contributing to the collection point signal. Where that information is available, ELISA optical density or titer measurements may be extrapolated to a bulk copies per person measurement, in a way that is analogous to the handling of viral copy number data.^41^ Because the protocol is able to detect antibodies in frozen archived wastewater samples, it becomes possible to analyze the SARS-CoV-2 pandemic retrospectively and to develop such models while simultaneously testing their predictive value against historic incidence levels. However, there are limitations to this approach that will need to be overcome prior to doing so. First, more extensive experimentation will be required to establish the limits of detection in the protocol and optimize it for widespread use. Second, the accuracy of an ELISA-based time series is contingent on the rate of antigenic evolution. We revealed a stark contrast between anti-S and anti-N IgG in late 2021 that is likely a consequence of a change in sensitivity of the anti-S ELISA assay to the significant mutational changes that occurred in the spike antigen during that time.^42^ Regardless, such challenges are readily surmountable by assaying additional samples and also assessing the sensitivity of results to variant-specific ELISA assays for delta- and omicron-specific antibodies. Using such approaches, existing samples collected throughout the COVID-19 pandemic can be used to conduct historic longitudinal studies that simultaneously quantify concentrations of antibodies and integrate that information with viral load and case data for the corresponding sewershed population. Such an approach represents an exciting next step for determining the promise of antibody detection in wastewater.

A wastewater surveillance approach to antibody abundance would complement rather than replace traditional serology. Associating ELISA optical density or titer measurements to equivalent concentrations in serum is challenged by individual variation of the immune response^43^ and heterogeneity in the rate of fecal shedding of detectable antibodies that is likely to be similar to heterogeneity in the rate at which viral particles are shed through feces.^44,45^ Wastewater surveillance has the potential to fill gaps in geographic settings where population-wide serological studies are not feasible, and reveal vulnerable neighborhoods or populations with low or undetectable antibody levels. Such an approach may be particularly useful for identifying and mitigating geographic health disparities. In particular, vaccine-mediated immunity hinges on prompt boosting with updated vaccines that reflect circulating variants.^13^ Currently rates of boosting in countries such as the United States are low and highly biased between populations.^46^ As predicting the local impact of the next SARS-CoV-2 infection wave requires an estimate of the durability of the immune response from earlier vaccination or natural infection, wastewater-based assessments could allow such data to be rapidly collected and thereby identify populations at risk.

## Methods

Prototype antibody assays first suggested the possibility of antibody detection from wastewater. 24 hour influent samples were collected from a South Burlington, Vermont wastewater treatment plant. 1L wastewater samples were homogenized and incubated with shaking at 4°C for one hour. The homogenate was clarified via centrifugation, 3500rpm at 4°C for 15 minutes, followed by 13000 rpm at 4°C for an additional 15 minutes. The clarified sample was then filtered through a 0.2um filter and run on a protein-G agarose column. Fractions were collected and subsequently, a Nanodrop spectrophotometer detected a peak indicating the presence of immunoglobulins, specifically IgG. This assay was not suitable for use on a large scale due to the volume of input material required and the high cost and limited availability of protein-G agarose column material to process 1L volumes. Building upon this observation, we developed a scalable protocol for quantification of IgG and IgA from wastewater using commercially available reagents.

### Sample collection and processing

Samples were collected from the study areas described in ^9^ and ^47^ following the study protocols described. Briefly, 24 hour composite samples were collected using ISCO portable autosamplers. For this study, three sites representative of typical wastewater surveillance sampling areas were selected. One of these is a high-traffic single building, the UNC Charlotte Student Union. The second site (Greenway) is a trunkline manhole with a sub-sewershed collection area that encompasses the entire UNC Charlotte campus. The third site is the Mallard Creek wastewater treatment facility, which is the Charlotte Water treatment plant serving a larger area that includes the university. Samples were either processed fresh or archived following collection in several 50mL conical tubes and stored at −80°C.

80 mL of wastewater was split into 2, 50mL conical tubes and centrifuged for 10 minutes at 10,000 x G at 20°C. Remaining supernatant liquid from centrifuged 50mL conicals was poured carefully into 2, new 50mL conical tubes, making sure not to pour solid pellet/particles along with supernatant. Sample supernatants can be stored at −80° C until ready for filtration. 15mL of liquid sample supernatant was poured into each 30kD filtration tube (Millipore) and centrifuged for 60 minutes at 4000 x G at 4° C. This is repeated for the entire sample to ensure sufficient concentration. Concentrated protein supernatant was removed from filtration tubes and stored at −80°C.

### ELISA detection of specific SARS-CoV-2 IgG and IgA

For ELISA detection of total human IgG and total human IgA, plates were pre-coated with specific antigen to human IgG by the manufacturer (Abcam), and the developer specifications were followed without modification. Briefly, a sandwich ELISA was performed using an antibody cocktail containing both capture and detector antibodies after addition of either neat or concentrated wastewater to the plate. The detector antibody is conjugated to horseradish peroxidase (HRP). After a 60 minute incubation with shaking at room temperature, the plates were washed thrice with proprietary wash buffer followed by the addition of the substrate tetramethylbenzidine (TMB) for 10 minutes. TMB detected HRP activity on bound antibodies. After 10 minutes, proprietary stop solution was added to stop the activity of TMB and change the pigment from blue (620 nm) to yellow (450 nm). After the addition of the stop solution, the plates were immediately read on a fluorescence plate reader (Biotek Synergy LX).

For ELISA detection of SARS-CoV-2 S protein-specific and N-protein specific and IgG, plates were pre-treated with SARS-CoV-2 S1 RBD protein by the manufacturer (Abcam), and the developer specifications were followed without modification. Briefly, concentrated wastewater was added to the pre-coated plates, and washed four times with proprietary wash buffer after 60 minutes of incubation with shaking. Biotinylated anti-human IgG was added post wash for 30 minutes with shaking and washed four times after incubation with wash buffer. HRP-conjugated streptavidin was added for 30 minutes and washed as above. Streptavidin has a high binding affinity to biotin, which was coupled to the anti-human IgG. After washing away excess HRP-streptavidin, TMB was added to the plate for 15 minutes to activate the HRP. After 15 minutes, proprietary stop solution was added to terminate the HRP/TMB reaction and the plate was read at 450 nm on a fluorescence plate reader (Biotek Synergy LX). A second set of assays was run using Abcam and Raybiotech ELISA products on the same set of input samples to ensure that the results were not platform dependent (Figure S1, Table S1). the Raybiotech assay was followed to the developer specifications without modification.

## Data Availability

Data produced and protocols used in the present study are included in full in the article. Authors will respond to any reasonable request for clarification or further information.

## Supplementary Information

**Figure S1:**
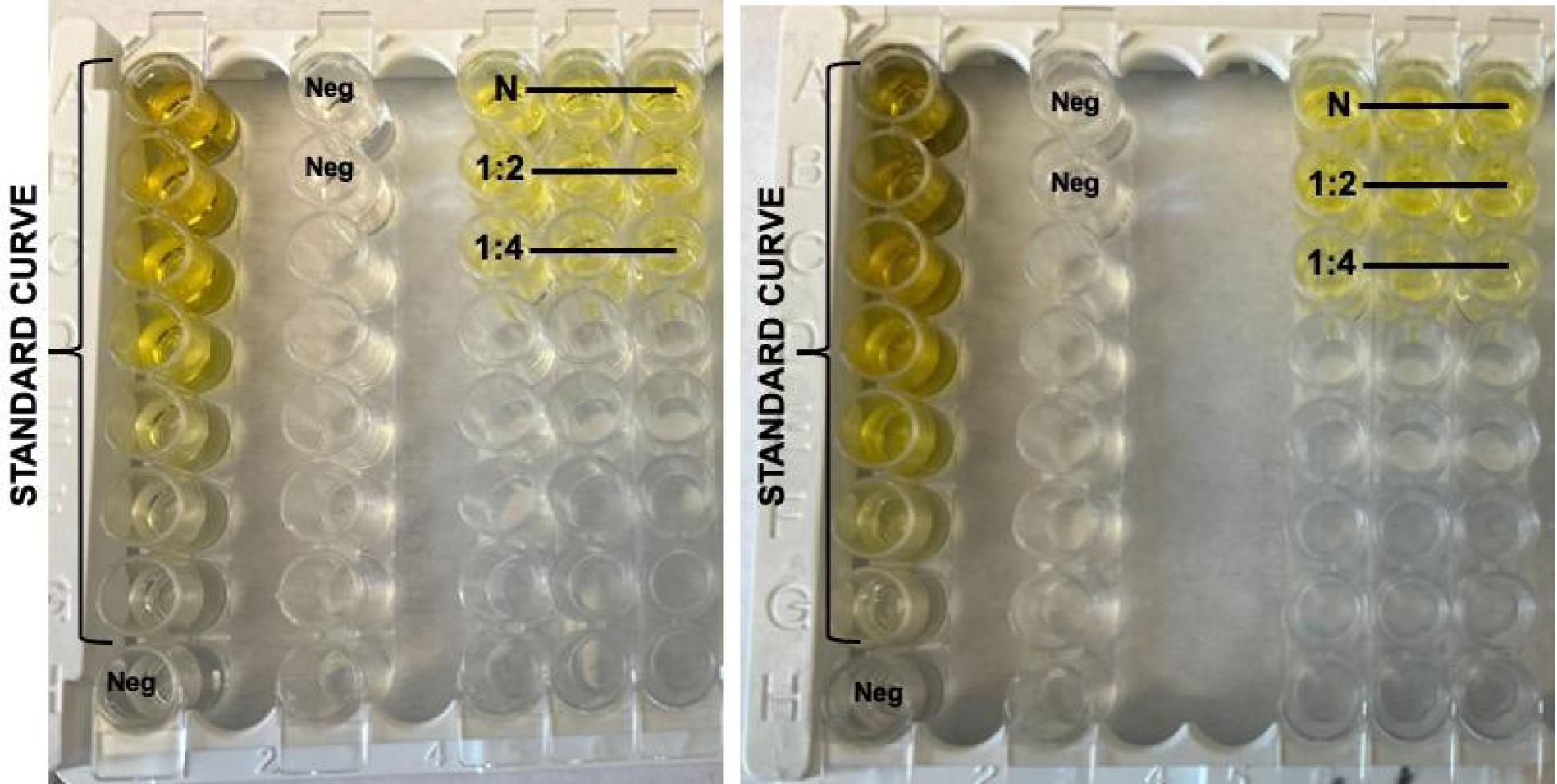
Concentrated wastewater evaluated for anti-SARS-Cov-2 S-Protein IgG by ELISA using Abcam (left) and Raybiotech (right) kits.

**Figure S2:**
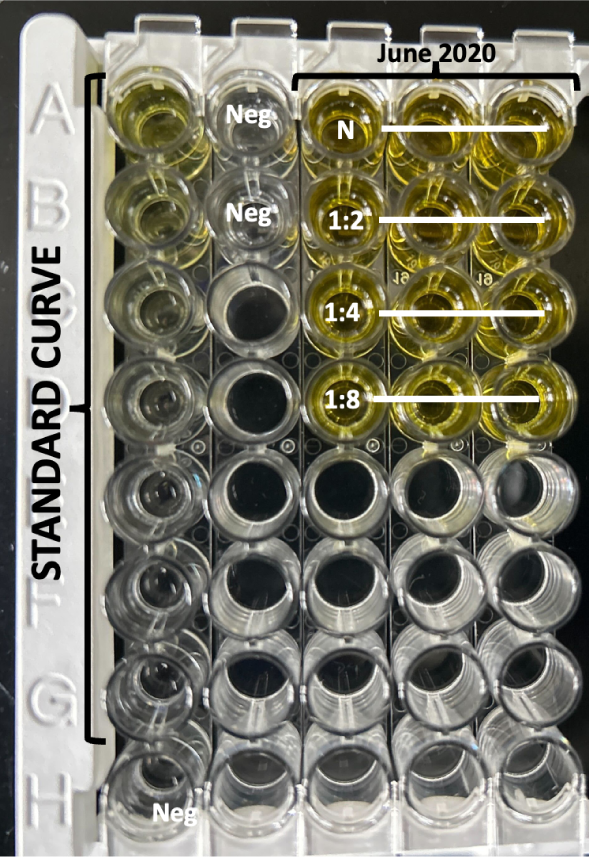
Concentrated wastewater from June 2020 evaluated for non-specific human IgG

**Figure S3:**
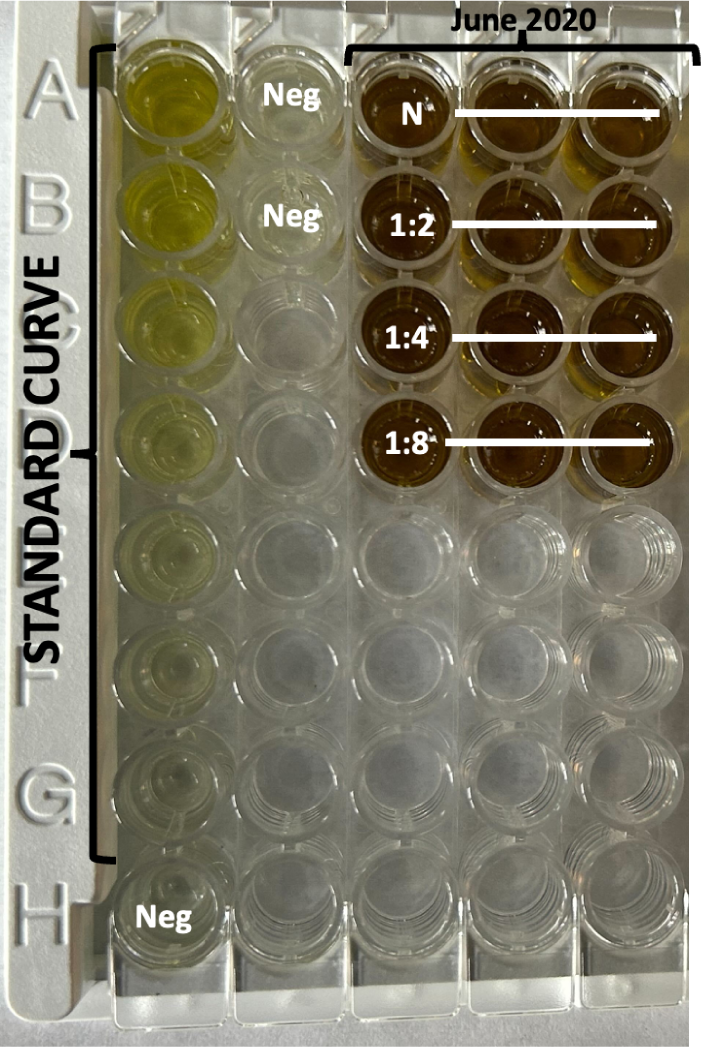
Concentrated wastewater from June 2020 evaluated for non-specific human IgA

**Table S1:** Average triplicate OD450 readings for COVID-19 S-Protein (S1RBD) Human IgG from Union wastewater collection site:

| Dilution | Abcam | Raybiotech |
| --- | --- | --- |
| Neat | 0.598 | 0.658 |
| 1:2 | 0.554 | 0.638 |
| 1:4 | 0.363 | 0.33 |
| Negative Control | 0.050 | 0.047 |

\*NB - per manufacturer's specifications, any value greater than twice the negative control is deemed positive.

**Table S2:** Average triplicate readings for non-specific human IgG in campus wastewater in June 2020.

| Dilution Factor | Average OD450 |
| --- | --- |
| Neat | 3.832 |
| 1:2 | > 4.000 |
| 1:4 | 3.392 |
| 1:8 | 2.157 |

\*NB - per manufacturer's specifications, any value greater than twice the negative control is deemed positive. Average Negative Control OD450: 0.051

**Table S3:** Average triplicate readings for non-specific human IgA in campus wastewater in June 2020.

| Dilution Factor | Average OD450 |
| --- | --- |
| Neat | > 4.000 |
| 1:2 | > 4.000 |
| 1:4 | > 4.000 |
| 1:8 | > 4.000 |

\*NB - per manufacturer's specifications, any value greater than twice the negative control is deemed positive. Average Negative Control OD450: 0.119

## Notes

### Competing Interest Statement

The authors have declared no competing interest.

### Funding Statement

Funding was provided by the UNC-Charlotte Division of Research.

### Summary of Updates

Additional data including total antibody assays of pre-pandemic samples and a time course spanning delta and omicron waves have been added.

